# Routine surveillance of kidney allograft rejection using a fully automated urinary CXCL9 and CXCL10 immunoassay

**DOI:** 10.1101/2023.07.26.23293106

**Authors:** Claire Tinel, Virginia Sauvaget, Laïla Aouni, Baptiste Lamarthée, Charlotte Leclaire, Christophe Legendre, Pierre Marquet, Corinne Normand, Marion Rabant, Fabiola Terzi, Dany Anglicheau

## Abstract

**Background:** For kidney transplant recipients, assessing non-invasively the individualized risk of acute rejection is one of the most unmet need. Urinary chemokines are one of the short-term most promising biomarkers, because of their simple and low-cost analytical method in easily accessible samples, and their high diagnostic performance consistently assessed over the last decade. In this study, we aimed at confronting all practical issues of routine implementation of kidney allograft rejection monitoring using urine CXCL9 and CXCL10.

**Methods:** The next-generation immunoassay Ella® was investigated as feasible quantification platform, from sample collection to render of the results. Urine CXCL9/10 levels were measured using Ella® microfluidic cartridges, across preanalytical (sample preparation, storage conditions, freeze-thaw cycles) and analytical (linearity, ranges, intra/inter-assay variability) performances studies, and diagnostic accuracy was assessed in comparison to the ELISA reference method, in urine samples from previously published cohorts.

**Results:** Upon assay preparation, Ella® appeared very efficient with a minimal workflow (urine sample thaw> centrifugation>1:2 dilution>loading) and a time to result of 90 minutes. Preanalytical studies showed high stability of urine CXCL9/10 levels across various temperatures (4° vs 25°C) and time (24/48/72H) before storage and over 5 freeze-thaw cycles. How complex urine matrix, analytical studies confirmed excellent linearity of dosage, as well as intra-assay (≤5%) and inter-assay precision. Across 1024 samples, Ella® results were highly correlated to those quantified by ELISA (P<0.0001), and further entered into our previously validated urine chemokine model. 268 out of the 1024 samples were collected at time of acute rejection (26.2%). In this cohort, accuracy was 0.85 for acute rejection diagnosis.

**Conclusion:** Ella® fulfils all prerequisites for clinical implementation of urinary chemokines monitoring. It has proven a robust, easy-to-use platform with unprecedented validation to quantify urine chemokines.

## INTRODUCTION

The standard of care for monitoring kidney transplant recipients has remained largely unchanged over the years, mostly relying on serum creatinine, proteinuria and anti-human leucocyte antigen (HLA) donor-specific antibodies (DSA) assessment. Any alarming modification of one or more of these parameters, prompts the decision to perform an allograft biopsy to assess the presence of an ongoing alloimmune injury. The histological evaluation, along with Banff classification, remains our most invaluable tool for routine surveillance, but comes with risk of bleeding, high cost, and requires trained pathologists. Therefore, addressing the shortcomings of current monitoring and developing diagnostic and prognostic biomarkers has been the aim of many studies^1, 2^.

In this drive toward innovative biomarkers, two main lines emerges: research use only investigations and strategies to develop clinical applications. The former often relies on high-throughput techniques using various omics approaches, and has provided novel insights and exciting perspective in disease pathways^3–5^. However, the complexity of these technologies, their cost and the sample selection for quality control along the process, makes it less fitted for routine work-up in the near future. The latter, takes advantage of minimally- to noninvasive body fluid collection, and focuses on clinical utility and practical implementation. In this respect, urine can be regarded as an ideal fluid for kidney allograft assessment, at the same time body waste and condensed reflection of intragraft injuries^6^.

The use of urine chemoattractant cytokines (chemokines) levels as a sum of all ongoing inflammatory processes within the allograft has been proposed, and further validated in several studies from independent research groups^7–12^. Among them, CXCL9 and CXCL10 have been identified as robust biomarkers that can be measured at the messenger RNA^10, 12^ or protein level^7–9, 11^. Urinary CXCL9 (uCXCL9) and uCXCL10 essentially appear to have a very high negative predictive value (NPV) for the diagnosis of ongoing and short-term acute rejection (AR)^13, 14^. Recently, we have refined the confounding factors in using urinary chemokines as non-invasive markers of AR. As a first, we have built a multiparametric fully-validated model, including uCXCL9/10 and their confounders, which proved its clinical utility^15^. Besides, we have investigated uCXCL10 as predictive biomarker in the course of BK virus (BKV) viremia, and we have showed that increased levels were associated with a significant decline in allograft function and an increased occurrence in antibody-mediated rejection (ABMR)^16^.

While urinary chemokines come to a high level of validation in experimental studies in research labs, clinical implementation requires development of robust, fast, reliable and affordable technics. Our previous experience highlighted how challenging immunological testing in urine sample can be. Hence, comparison of several ELISA kits demonstrated that urinary CXCL9 levels can only be quantified by a kit necessitating overnight plate precoating, which hinders transfer to routine (**Supplemental Figure 1**).

To overcome these issues, we investigated the next-generation immunoassay Ella® (ProteinSimple™), considering its alleged ease of use, reliability on urine samples and rapidity in result rendering. The Ella® method, relies on an immunoassay quantification, using cartridges of different formats, which is claimed to have a superior workflow and a higher throughput and quality than ELISA. Each well of the cartridge is sampled three times to give results in triplicate. Each well actually corresponds to a microfluidic channel, directly coated with the capture antibody. The antibodies as well as the fluorescent detector are pre-functionalized in the cartridge, with a built-in standard curve. Of note, some Ella® assays have been tested and validated for quantification from human urine samples, but not CXCL9 and CXCL10 cartridges.

In this study, we evaluated the Ella® platform as a feasible technique for routine implementation of urine chemokine monitoring after kidney transplantation, and validated each workflow step from sample collection to results reporting. More precisely, we investigated preanalytical sample processing from collection to storage and Ella® analytical performance from urine sample, as to provide suggestions of standard operating procedures, an essential factor in ensuring excellent sample quality and reliable results^17^. For clinical validation, we assessed whether the Ella® platform provided close and useful results in comparison to the reference ELISA technique.

## MATERIALS AND METHODS

### Study samples and cohorts

Samples for Ella® technical validation (preanalytical and analytical performance studies) were all taken from the local Transplantation Biobank (DC-2009-955, Necker Hospital, Paris, France). Urine samples were routinely collected from kidney transplant recipients as part of transplant care, with exception for the “storage study” where freshly emitted urine samples (N=12) were prospectively collected. Besides, urine samples (N=10) were collected from kidney living-donors (KD) at the time of their pre-donation evaluation.

Samples for the clinical validation study belong to previously published cohorts and comprised N=275 samples for Cohort A^11^ , N=37 samples for Cohort B^15^, N=147 samples for Cohort C^15^ and N=330 samples for cohort D^15^. Ethics Committee of Ile-de-France XI (#13016) gave ethical approval for this work and all participating patients provided written informed consent.

### ELISA methods

We previously described the extensive protocols for urinary chemokines quantification by enzyme-linked immunosorbent assay (ELISA)^15^. Briefly, uCXCL9 was measured using Human CXCL9/MIG DuoSet ELISA kit (Bio-Techne, Minneapolis, USA), with a protocol optimized for quantification from a urine sample. Human CXCL10/IP10 Quantikine ELISA Kit (Bio-Techne,) was used according to the manufacturer’s instructions. Optical densities were measured using a Multiskan FC plate reader (Thermo Fisher, Illkirch, France). All measurements were performed in duplicate.

### Ella® immunoassay methods

Urine CXCL9 and CXL10 levels were measured using the Ella® microfluidic Single Plex cartridges (ProteinSimple™, Bio-Techne, Noyal Châtillon sur Seiche, France), following the manufacturers’ instructions. Briefly, urine samples were stored frozen at -80°C, thawed on ice, then centrifugated at 1500 relative centrifugal force (g) for 2 minutes, as to pellet all debris which might cause microfluidic channel obstruction. For Single Plex cartridge loading, 50µL of each diluted urine supernatant sample (1:1 in Sample Diluent) or quality control was added to the wells, as well as 1 mL of Wash Buffer in the dedicated inlet. The automated Ella® immunoassay protocol was then initiated, including automated three times sampling of each well to give results in triplicate. Measurement of creatinine was performed in the same urine samples using the Creatinine Parameter Assay Kit (Bio-Techne).

### Sample preparation study

As part of the local routine biobanking (reference method here), urine samples are collected and processed as follows: samples are kept at room temperature (RT) until centrifuged at 3300 rpm for 20 minutes at 4°C within 3 hours of collection. The supernatant is collected and stored with or without protease inhibitors (cOmplete™, Roche Diagnostics, Meylan, France) at -80°C. In this Sample Preparation Study, both aliquots of 25 urine samples were thawed on ice and urinary chemokines were quantified by Ella® technique in a single batch. Chemokine levels in each sample were compared according to the addition or not of protease inhibitors during sample preparation.

### Storage study

Fresh urine samples (N=5) were prospectively collected from hospitalized kidney transplant recipients presenting with a condition usually associated with high urinary chemokines levels (*i.e.* AR, BKV replication or bacterial urinary tract infection), and split into 7 aliquots subjected to various procedures to produce 7 samples from each. A first aliquot (standard tube) was immediately centrifuged and stored without protease inhibitors at -80°C. The other aliquots were left for 24/48/72 hours, respectively at 4°C or at room temperature (RT). Samples were centrifuged immediately before storage without protease inhibitors, and kept at -80°C until analysis by Ella® technique in a single batch. Chemokine concentrations in each sample type were compared to those from the corresponding standard tube.

### Freeze/thaw cycles study

Urine samples (N=5) were aliquoted into 5 tubes and stored at -80°C without protease inhibitors until further analysis. Samples were thawed on ice during 2h and frozen again at - 80°C on consecutive days. This procedure was performed in respective aliquots once (T1), twice (T2), three (T3), four (T4) or five (T5) times. Chemokine concentrations in T2/T3/T4/T5 sample were compared to those from the matching T1 tube (see details in *Statistical analysis*). In this study, T1 corresponding to one freeze-thaw cycle is considered as the reference method and mirrors clinical use where samples are usually stored until filling-up the assay-plate.

### Linearity

Urine samples (N=10) with a previous chemokine quantification were chosen to encounter for a broad range of CXCL9 and CXCL10 concentration, and diluted 1:2, 1:4, 1:8 and 1:16 in Sample Diluent (SD13, Simple Plex™, Bio-Techne). All diluted samples were assayed within a single run. Linearity was assessed by mean of the coefficient of variation (CV) with 1:2 dilution taken as the reference sample, and by Spearman correlation tests.

### Accuracy and limit of quantification

Accuracy of Ella® internal calibration curve on urine samples was assessed by using two recombinant Human CXCL9 proteins from two different manufacturers. Human CXCL9/MIG DuoSet ELISA kit (Bio-Techne) was reconstituted with 0.5mL of Reagent Diluent (RD, Catalog # DY995, Bio-Techne) and Recombinant Human CXCL9 (Biolegend, San Diego, USA) was reconstituted with 50μL of PBS. Those recombinant CXCL9 proteins were further diluted in either in Sample Diluent SD13 (Simple Plex™, Bio-Techne) used for Ella® procedure or in pooled urine samples from KD. Urinary CXCL9 and CXCL10 levels were hence measured in urine samples (N=10) collected from KD prior to kidney donation. Samples from various age, male and female KD were selected to bring diversity (**Supplemental Table 1**) and pooled together.

For each of the diluent, eleven points standard curve using 2-fold serial dilutions was prepared. The resultant samples were quantified both by Ella® in a single batch. Percent recovery was calculated for each of the 11 points, based on the found concentration and the theoretical concentration.

### Within- and between-run precision

Within-run (intra-assay) precision was assessed on urine samples (N=5) quantified twice on a same CXCL9 or CXCL10 cartridge. Between-run (inter-assay) variation was assessed for CXCL9 and CXCL10 on urine samples (N=5) quantified by two different technicians on different days with cartridges from the same lot. For CXCL9, intermediate precision was further refined in a larger number of samples (N=32) to assess technician-to-technician and day-to-day variations in high, mid and low CXCL9 concentration samples. Precision was expressed as CV.

### Clinical accuracy study

To evaluate the diagnostic performances of urinary CXCL9 and CXCL10 quantified by Ella® as compared to the reference ELISA method, we used all available urine samples from four previously published cohorts^11, 15^. Among the 1170 urine samples included in the original works, enough material was available for 1124 of them (96.1%). ELISA and Ella® results were compared using Spearman correlation tests and Bland-Altman analysis. These results were further used to investigate how the modification in quantification method might influence the performance of our 8-parameter chemokine model. All samples with no missing data for all 8 variables (N=1024) were used to build several risk prediction models using logistic regression and random forest analyses. Accuracy was assessed by mean of an Area Under the recipient operating Curve (AUC).

### Statistical analyses

Patient and donor characteristics are described by numbers, percentages and frequencies for categorical variables. We report continuous variables using means and standard deviations (SD) or medians and interquartile ranges (IQR) for variables with a skewed distribution. Changes in concentrations of chemokines over time, temperature or freeze-thaw cycles were analyzed using one-way repeated measures analysis of variance (RM-ANOVA) followed by Sidak’s multiple comparisons tests. AUC’s were compared with the Delong’s test. Statistical analyses were performed using Graphpad Prism version 9.0.1 (GraphPad Software, San Diego, USA) and with R software (R Development Core Team, R version 4.0.3 and R studio version 1.3.1093).

## RESULTS

### Effects of preanalytical sample processing on urine chemokines assessment

If urinary chemokines are to be used for routine surveillance of KTRs, urine sample collection and processing have to be optimized to fit hospital’s constraints (**Figure 1, Sample collection & storage**). In research, protease inhibitors (PI) are usually added after urine supernatant collection to prevent protein degradation upon long-term storage. However, this additional step during sample preparation is time consuming, costly, and might prevent consistent practice between centers. Thus, valid information about necessity of PI addition to prevent CXCL9 and CXCL10 degradation is essential. Chemokine levels of 25 urine samples were compared according to the addition or not of PI during sample preparation. Median time from sample collection to quantification was 147 days [IQR: 127-184]. **Figure 2A** shows a nearly perfect correlation for CXCL9 (Spearman r=0.98 [95% CI: 0.96-0.99], P<0.0001), with a mean with/without PI ratio=1, as well as for CXCL10 (Spearman r=0.96 [95% CI: 0.92-0.98], P<0.0001), with a mean with/without PI ratio=1.1.

**Figure 1:**
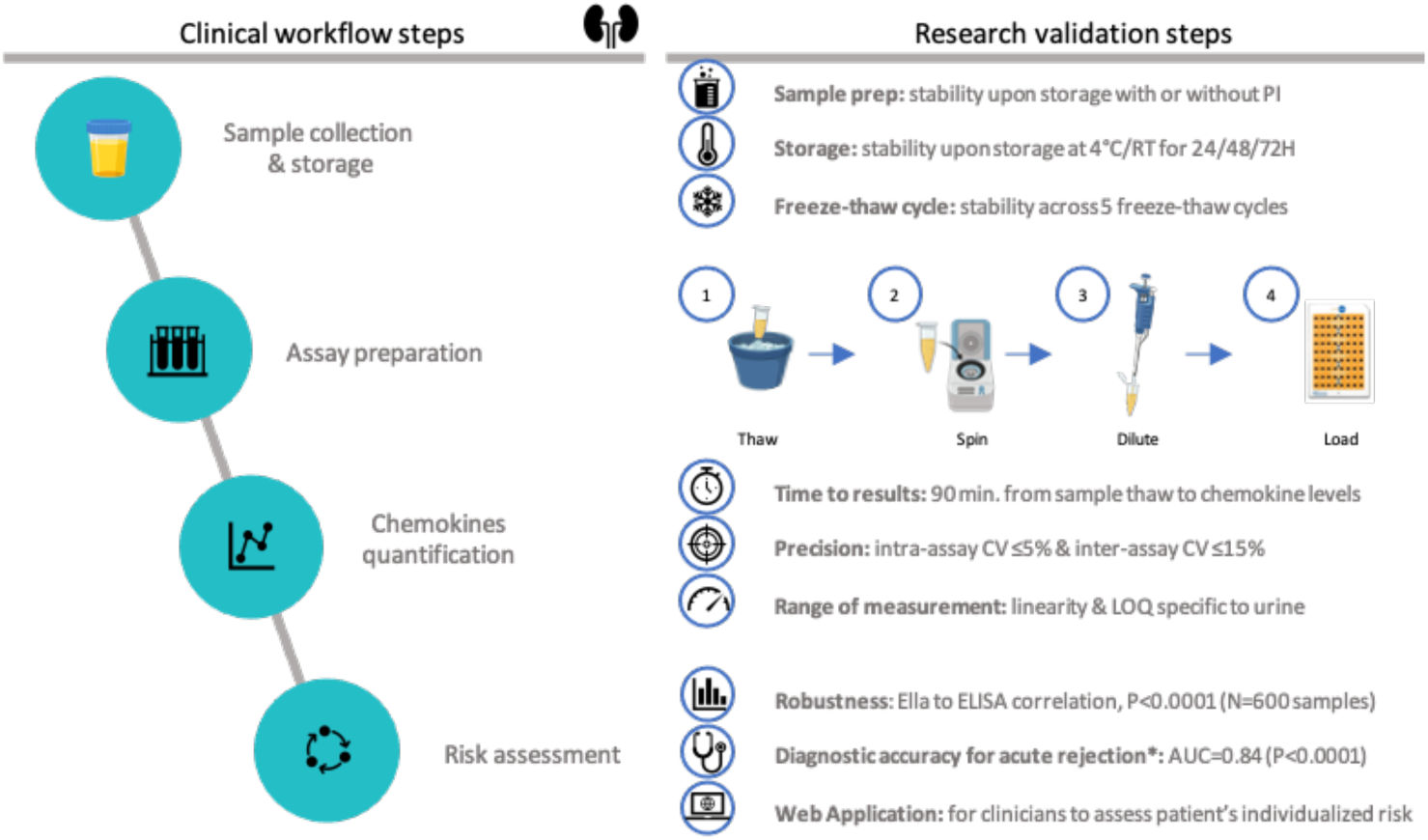
Overview on clinical workflow steps and research validation studies performed to assess Ella® quantification of urine chemokines: bringing research innovation to daily care. **Left panel** illustrates the 4-step workflow if urine chemokine were used in daily practice: sample collection and storage, assay preparation, chemokines quantification and acute rejection risk assessment. For each clinical step, the **middle panel** shows the corresponding validation studies which were conducted, independently from manufacturer’s certification. The **right panel** presents the web application that was developed to prompt the use of urine chemokines in daily practice. A screen shot of the user interface is shown, illustrating how clinical and biological data can easily be filled in. Abbreviations: AUC, area under the curve; CV, coefficient of variation; LOQ, limit of quantification; min, minutes; prep, preparation; PI, protease inhibitors; RT, room temperature.

**Figure 2:**
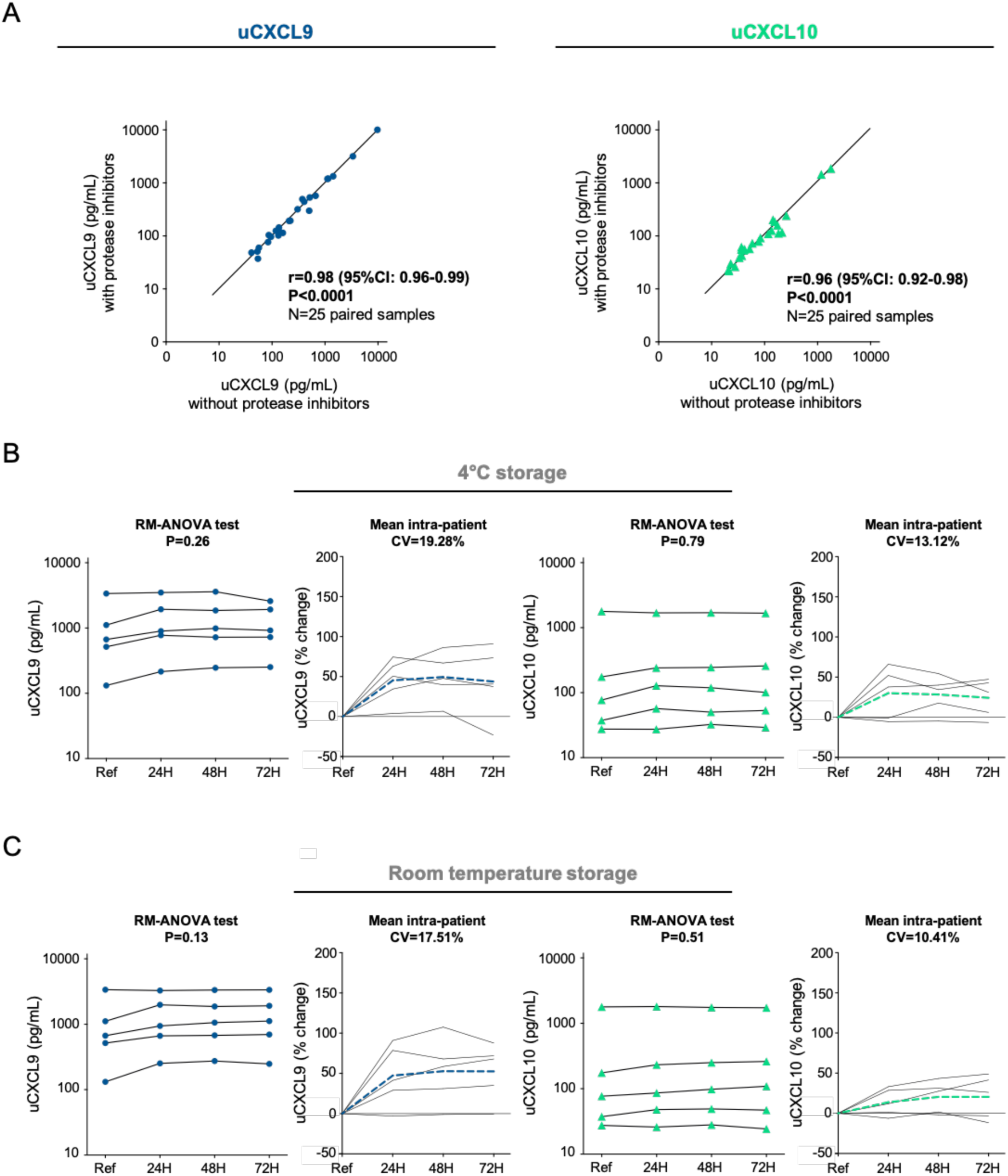
Effects of preanalytical sample processing on urine chemokine assessment. **A. Sample preparation study.** Correlation plots illustrating relationships between uCXCL9 (left panel) and uCXCL10 (right panel) measurement from 25 pairs of urine samples, prepared and stored with or without protease inhibitors. P-value and r from a Spearman correlation test. **B-C. Sample storage study**. Changes in urinary chemokine concentration over various processing delay (24H, 48H and 72H) and at 4°C (**Panel B**) or room temperature (**Panel C**) storage conditions. Raw data from 5 individual samples are presented as solid lines for CXCL9 (navy blue dots) and for CXCL10 (green triangles). The percentage change was calculated at each timepoint in comparison to reference sample, and mean percentage change at each timepoint is presented by dashed lines (navy blue for CXCL9 and green for CXCL10). RM-ANOVA was performed to assess intra-patient variability. CV was calculated between all 4 urinary chemokine levels and mean CV is given as the mean of all intra-patient CVs. Baseline refers to standard sample processing (within 3H after collection) and storage (freezing at -80°C).

### Effects of processing delay and storage conditions on urine chemokines assessment

If routinely implemented, urine chemokine assessment might not be available in each single hospital and shipment to a centralized reference center might be considered. Besides, freezing a urine specimen prior to centrifugation may cause cell lysis upon thawing, allowing cellular cytoplasmic protein to contaminate the urine specimen. In research, an early centrifugation step is thus usually performed to pellet cells, but it requires an available technician and a dedicated equipment. Thus, we investigated the influence of time and storage conditions on chemokine quantification. Fresh urine samples from 5 patients were kept at 4°C or RT for respectively 24, 48 or 72 hours. Centrifugation to pellet urine cells and collect urine supernatant was performed immediately before -80°C storage.

Within-person stability assessment showed similar results of CXCL9 and CXCL10 levels over time for samples kept at 4°C (P=0.26 and P=0.79, **Figure 2B**) or at RT (P=0.13 and P=0.51, (**Figure 2C**). Up to 72 hours at RT, mean intra-patient CV did not exceed 20% for CXCL9 (4°C, CV=19.28%; RT, CV=17.51%) and 15% for CXCL10 (4°C, CV=13.12%; RT, CV=10.41%). Percent change in chemokine level was consistently positive across conditions, indicating a minor increase in chemokine levels upon time. Main variation happened within the first 24 hours, suggesting cell lysis from urine cell pellet with adds-on from intracellular chemokines.

### Effects of repeated freeze–thaw cycles on urine chemokines assessment

Next, we investigated the influence of repeated freeze–thaw cycles on both chemokines quantified in 5 urine samples (**Table 1**). In comparison to samples thawed only once (T1), up to 4 additional cycles (T2-T5) did not significantly change within-patient levels of CXCL9 (P=0.79) and CXCL10 (P=0.26). The percentage change and CV in CXCL9 and CXCL10 concentration were calculated for each refrozen sample in comparison to the baseline T1 sample. With mean CV of 10.63% for CXCL9 and 18.90% for CXCL10, both assays were found to meet the FDA acceptance criteria for bioanalytical method validation (<20%, https://www.fda.gov/files/drugs/published/Bioanalytical-Method-Validation-Guidance-for-Industry.pdf). Percent change remained low for both chemokines and was consistently negative for CXCL10, suggesting a more fragile protein, with possible degradation over repeated freeze-thaw cycles.

**Table 1:** Changes in urinary chemokine concentration (raw value and percentage of baseline) over 5 freeze-thaw cycles.

| uCXCL9 |  |  |  |  |  |  |  |
| --- | --- | --- | --- | --- | --- | --- | --- |
| Sample ID | Reference value (pg/mL) | Value (% change vs T1) |  |  |  | Mean % change | Mean CV (%) |
|  | T1 ± SD | T2 | T3 | T4 | T5 |  |  |
| S1 | 41 ± 4.1 | 38.3 (-6.6) | 30.4 (-25.9) | 39.4 (-3.9) | 37.4 (-8.8) | -11.28 | 9.42 |
| S2 | 87.2 ± 6.8 | 80.9 (-7.2) | 90.3 (3.6) | 98.1 (12.5) | 82.9 (-4.9) | 0.97 | 6.90 |
| S3 | 223 ± 20.0 | 242 (8.5) | 219 (-1.8) | 269 (20.6) | 232 (4.0) | 7.85 | 8.83 |
| S4 | 503 ± 41.4 | 438 (-12.9) | 466 (-7.4) | 392 (-22.1) | 468 (-7.0) | -12.33 | 11.82 |
| S5 | 376 ± 43.5 | 468 (24.5) | 436 (16.0) | 482 (28.2) | 407 (8.2) | 19.22 | 16.18 |
| Mean | - | - | - | - | - | **0.89** | **10.63** |

| uCXCL10 |  |  |  |  |  |  |  |
| --- | --- | --- | --- | --- | --- | --- | --- |
| Sample ID | Reference value (pg/mL) | Value (% change vs T1) |  |  |  | Mean % change | Mean CV (%) |
|  | T1 ± SD | T2 | T3 | T4 | T5 |  |  |
| S1 | 20.9 ± 4.53 | 14.8 (-29.2) | 10.6 (-49.3) | 12.9 (-38.3) | 9.35 (-55.3) | -43,00 | 41.75 |
| S2 | 40.5 ± 5.87 | 26.3 (-35.1) | 30.5 (-24.7) | 38 (-6.2) | 30.6 (-24.4) | -22.59 | 19.51 |
| S3 | 117 ± 4.30 | 109 (-6.8) | 107 (-8.6) | 114 (-2.6) | 108 (-7.7) | -6.41 | 5.13 |
| S4 | 214 ± 27.01 | 162 (-24.3) | 163 (-23.8) | 141 (-34.1) | 176 (-17.8) | -25,00 | 22.83 |
| S5 | 155 ± 9.56 | 153 (-1.3) | 149 (-3.9) | 168 (8.4) | 142 (-8.4) | -1.29 | 5.27 |
| Mean | - | - | - | - | - | **-19.66** | **18.90** |

Abbreviations: CV, coefficient of variation; ID, identity; SD, standard deviation.

### Linearity and range of measurement

CXCL9 and CXCL10 Ella® cartridges have not been validated on human urine samples. Considering the wide range of pH and urine specific gravity, and that urine complex matrix may hinder immunologic testing, we run an in-house validation of all aspects of analytical performance of the assay (**Figure 1, Chemokines quantification**). Ella® cartridges are provided with an internal calibration curve, *i.e.* a relationship between fluorescence and known concentrations of the analyte. But a calibration curve should be prepared in the same biological matrix as the sample. First, we investigated the ability of the assay to produce results that are directly proportional to the concentration of analyte in the urine sample. Linearity was assessed from 10 urine samples with a broad range of chemokine values from previous measurement, subjected to serial dilution (1:2, 1:4, 1:8 and 1:16). A high repeatability for each sample was assessed with mean intra-patient CV of 10.2% for CXCL9 and 9.3% for CXCL10 (**Figure 3A**). From a Spearman correlation analysis between each dilution factor, all r values were ≥ 0.98 (**Figure 3B**). For CXCL10, linearity was confirmed within the complete range given by the manufacturer (dilution-corrected range 1.2-1840 pg/mL). For CXCL9 (manufacturer’s range: 39.8-60,800 pg/mL), linearity was found reliable between 100 and 10,000 pg/mL, but was less clear for extreme values. Hence for sample 10 (**Figure 3A**), CXCL9 deviated from 8732 pg/mL (1:8 dilution) to 17464 pg/mL (1:16 dilution, CV=47.1%, %change=100). To further define CXCL9 lower and upper limits of quantification (LLOQ-ULOQ) on urine, we used recombinant CXCL9 serially diluted (1:2) into Sample Diluent or into pooled urine from healthy kidney donors, all with undetectable CXCL9 levels (**Supplemental Table 1**). Recovery at 11 different spiked concentrations (N=3) showed less reliable CXCL9 assessment below (expected value) 31.3 pg/mL and above 2000 pg/mL (**Table 2**). Overall, our linearity and recovery data support the following LLOQ and ULOQ on urine sample: 39.8-2000 pg/mL (CXCL9) and 0.6-920 pg/mL (CXCL10).

**Figure 3:**
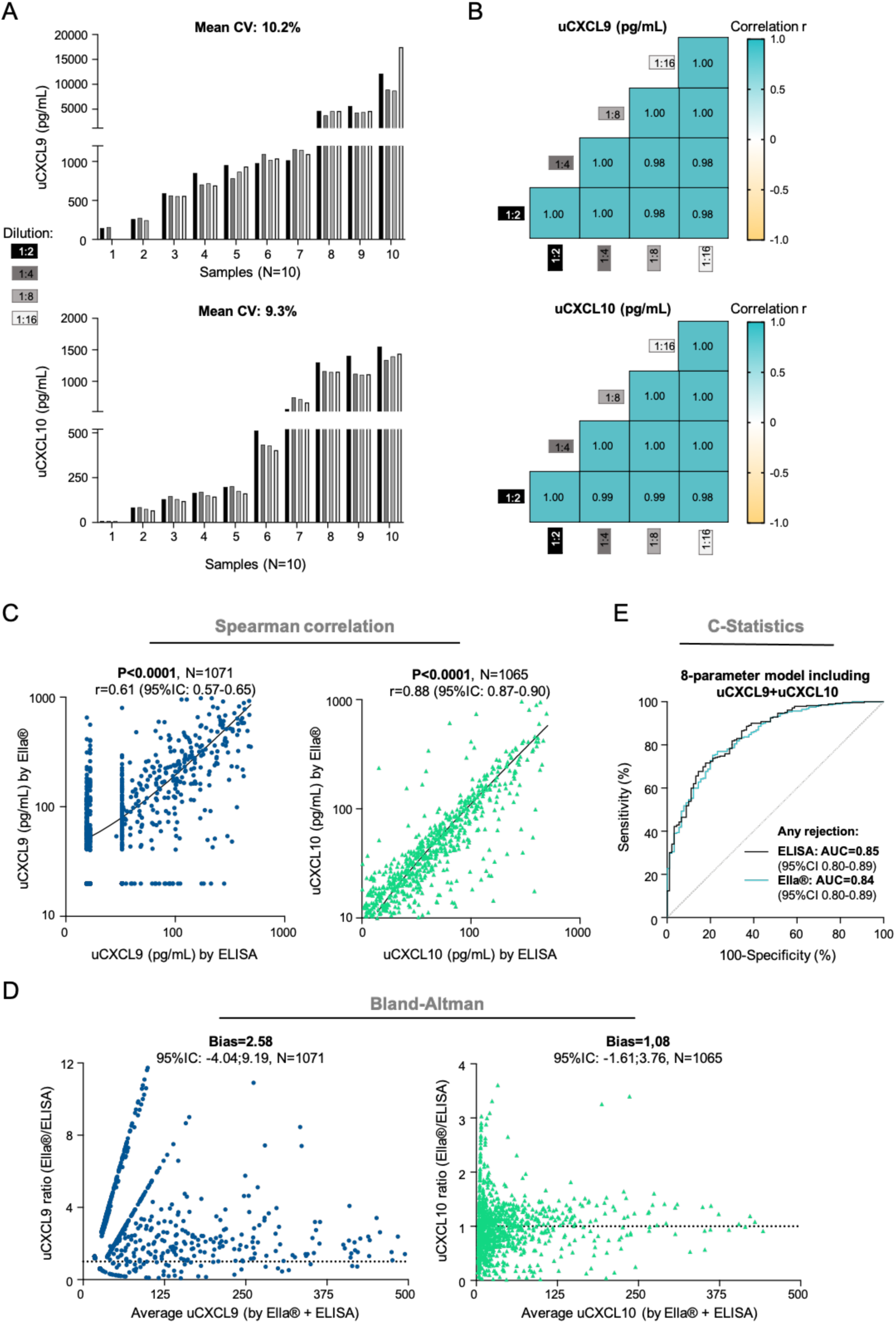
In-house validation of Ella® assay performance for chemokine quantification from a urine sample. **A-B. Linearity study. A.** Histograms showing uCXCL9 (upper) and uCXCL10 (lower) levels from 10 KTRs urine samples, serially diluted (1:2, 1:4, 1:8 and 1:16) before quantification. For each patient, CV was calculated between each diluted sample and the reference sample (1:2 dilution), and mean CV is given as the mean of all intra-patient CVs. **B.** Correlogram illustrating correlation between uCXCL9 (upper) and uCXCL10 (lower) levels after serial dilution for the same 10 KTRs. R values from a Spearman correlation, all P-values <0.0001. **C-D. ELISA to Ella**® **correlation study**. **C.** Correlations between uCXCL9 (N=1071, navy blue dots) and uCXCL10 (N=1071, green triangles) levels when measured by ELISA (x-axis) or by Ella® (y-axis) methods. R values from a Spearman correlation, all P-values <0.0001. **D.** Bland-Altman analysis. For each chemokine, Ella®/ELISA ratio (y-axis) is plotting against average chemokine levels (x-axis). Ella®/ELISA ratio of 1 would indicate perfectly superimposable values resulting from both techniques. Bias refers to deviation from this ideal ratio. For representation purpose, only average values <500pg/mL are shown. **E.** Diagnostic accuracy (C-statistics) of Ella® results tested against reference ELISA results. ROC curves illustrating the diagnostic performance of the 8-parameter chemokine model for any acute rejection, when trained on Ella® or ELISA results.

**Table 2:** CXCL9 recovery study to validate urine-specific ranges of measurement.

| CXCL9 in urine matrix (N=3) |  |  |  |  |  |  |  |  |  |  |  |
| --- | --- | --- | --- | --- | --- | --- | --- | --- | --- | --- | --- |
| **Expected value**<br>(pg/mL) | 7.8 | 15.6 | 31.3 | 62.5 | 125 | 250 | 500 | 1000 | 2000 | 4000 | 8000 |
| **Mean values**<br>(pg/mL) | 22.6 | 24.7 | 32.7 | 54.7 | 105.8 | 245.7 | 537.0 | 1190.0 | 2571.7 | 6710 | 21462.3 |
| **Range**<br>(pg/mL) | 04-53 | 10-53 | 19-58 | 24-89 | 74-132 | 173-294 | 422-630 | 1030-1392 | 2399-2770 | 5193-9057 | 14368-29492 |
| **Avg% of expected** | 289.7 | 158.3 | 104.7 | 87.5 | 84.6 | 98.3 | 107.4 | 119.0 | 128.6 | 167.8 | 268.3 |
| **Range (%)** | 52-679 | 65-342 | 60-186 | 38-143 | 60-106 | 69-118 | 84-126 | 103-139 | 120-139 | 130-226 | 180-2369 |
| **Proposed LOQ** | LLOQ=39.8 pg/mL |  |  | In range |  |  |  |  |  | ULOQ=2000 pg/mL |  |

Expected values: spiked concentrations of recombinant CXCL9 diluted in pooled-urine samples from 3 healthy donors (N=3). CXCL9 values are presented as mean values and average percent of expected values with respective ranges. Abbreviations: Avg, average; CV, coefficient of variation, LLOQ, lower limit of quantification, ULOQ, upper limit of quantification.

### Within- and between-run precision

For both assays, precision was assessed on 5 urine samples quantified twice on the same cartridge (intra-assay precision), or quantified twice by different technicians on different days (inter-assay precision). The intra-assay and inter-assay CVs were 4.7% and 15.3%, respectively for CXCL9, and 2.6% and 16.6%, respectively for CXCL10 (**Table 3**). For CXCL9, intermediate precision was further refined in a larger number of samples (N=32) to assess technician-to-technician and day-to-day variations in high, mid and low CXCL9 concentration samples. Under the same set of conditions and within a short interval of time, repeatability ranged from 3.8% (mid CXCL9) to 11.6% (low CXCL9). When investigating the random error introduced by factors like specific technicians, between-run variation was also found acceptable with CV ranging from 9.6% (low CXCL9) to 15.3% (high CXCL9). The inter-assay CV for all 37 tested samples averaged 10.3%.

**Table 3:** Within- and between-run precision.

| Analyte | Intra-assay precision<sup>a</sup> |  | Inter-assay precision<sup>b</sup> |  |
| --- | --- | --- | --- | --- |
|  | Samples (N) | Mean CV (%) | Samples (N) | Mean CV (%) |
| CXCL9 | 5 | 4.7 | 5 | 15.3 |
| CXCL10 | 5 | 2.6 | 5 | 16.6 |
| **Inter-assay precision (CXCL9)** |  |  |  |  |
| Group | *Same technician; same lot* |  | *Different technicians; same lot* |  |
|  | Samples (N) | Mean CV (%) | Samples (N) | Mean CV (%) |
| Low CXCL9 (<100pg/mL) | 5 | 11.6 | 5 | 9.6 |
| Mid CXCL9 (100<CXCL9<300pg/mL) | 6 | 3.8 | 6 | 12.2 |
| High CXCL9 (CXCL9 >300pg/mL) | 5 | 7.5 | 10 | 15.3 |
| All samples | 16 | 7.4 | 21 | 13.1 |

Abbreviations: CV, coefficient of variation. <sup>a</sup>Samples quantified twice in triplicate on a same cartridge. <sup>b</sup> Samples quantified on different days by different technicians, cartridges from the same lot.

### Clinical accuracy study

To evaluate the clinical performances of uCXCL9 and uCXCL10 quantified by Ella®, over 1000 samples belonging to 4 previously published cohorts (Cohort A^11^, B-D^15^) were quantified again using the Ella® method. We observed a high degree of correlation between uCXCL9 and uCXCL10 measurements by Ella® and by the reference ELISA method (P<0.0001, **Figure 3C**). More specifically, assessments from the two methods were compared using Bland-Altman test (**Figure 3D**). For uCXCL10, both methods provided very superimposable values, with uCXCL10 Ella®/ELISA ratio mostly distributed around 1 (Bias=1.08; 95%CI, -1.61:3.76). For uCXCL9, though highly correlated, numerical values were found higher when quantified by Ella® in comparison to ELISA (Bias=2.58; 95%CI, -4.04:9.19). Combined with the previous recovery study using recombinant CXCL9, these results suggest that Ella® might provide a more accurate numerical quantification for CXCL9, than ELISA did. Finally, we previously established a model of AR risk using urine chemokines and their confounding factors. Using the same development cohort, we trained the model on Ella®-generated data, instead of ELISA data. Overall, discrimination performance of the 8-parameter model for the diagnostic of AR remained equivalent (AUC_Ella®_=0.8406, AUC_ELISA_=0.8481, DeLong’s P-value=0.49, **Figure 3E**).

### Assessment of acute rejection risk using urine chemokines

Our original model was derived in KTR from Necker Hospital, and validated in an external single-center cohort and in a prospective multicenter unselected cohort^15^. All samples from these 3 cohorts have since been quantified again by Ella® method, as well as all samples from our princeps study on urinary chemokines^18^, enabling to train and validate the model on a large set of Ella® data (N=1082 samples). The resulting 8-parameters model reached an AUC of 0.84 for any rejection diagnosis. For clinical assessment of a patient’s risk for AR, and either prompt the decision in performing a biopsy, either argue for avoiding a unnecessary biopsy, a web application calculator is under development in our lab. Transplant specialists would easily enter their patients clinical data (age, gender), serum lab tests (creatinine, DSA and BKV viral load) and urine lab tests (creatinine, uCXCL9 and uCXCL10 levels), and rapidly get an accurate risk prediction.

## DISCUSSION

In the present study, we aimed to concretize the use of well-established urine chemokines CXCL9 and CXCL10 to routine monitoring of KTRs, with 2 key points: the validation of an automated assay and the development of a web application for AR risk calculation. We used an Ella® measurement of CXCL9 and CXCL10, which have been validated for quantification from plasma, serum and cell culture supernatant but not from urine samples. Our assessment of the Ella® platform from KTR urine sample shows adequate performances with regard to reproducibility, validity, stability after several freeze-thaw cycles, and upon various storage conditions. Results were for the most part reproducible, with laboratory intra- and inter-assay CVs between 3 and 17%, thus meeting the FDA acceptance criteria for bioanalytical method validation^19^. Most importantly, Ella®-quantified chemokines showed similar performance in diagnostic accuracy for acute kidney allograft rejection than ELISA.

In collection and storage of biological samples for developing biomarkers, degradation upon processing and storage is an important issue^20, 21^, but consistent information on biomarkers stability is often lacking^22^. To avoid proteolysis, the use of protease inhibitors is widespread when using immunoassays where no interference was shown, as opposed to proteome analysis^23, 24^. However, its benefit remains unclear and more importantly, these additional step and cost might prevent an easy and standardized collection in the clinics. Moreover, adds-on tests are regularly requested from laboratory, leading to repeated freeze-thaw cycles from a single samples (if not previously aliquoted). The water-to-ice transformation during sample freezing is a well-studied process where time is important: in a slow-freezing process, large, and solid abrasive ice crystals are formed while in snap-freezing process (liquid nitrogen), ice crystals are small and amorphous, maintaining integrity of samples^25^. Because of the hazardousness of liquid nitrogen handling, snap-freezing is not feasible for routine storage of samples. Thus the question of how slow-freezing at -80°C impacts the urine samples and the immunoassay results remains. We therefore compared urine chemokine levels from paired samples with or without addition of PI, and up to 5 freeze-thaw cycles, and observed no significant difference in urine chemokine levels. We observed a small reduction in CXCL10 upon storage without PI and upon repeated freeze-thaw cycles, with degradation possibly explained by unfolding of the protein or by its cleavage by proteases. Moreover, transient storage at 4 °C or at RT is likely to affect the concentration of chemokines prior to mid-term storage at -80°C, whether by protein degradation or by release from intra-cellular compartment. To remove unnecessary components and to collect only pellet-free urine supernatant, a 20 minute-long centrifugation at 4°C is usually immediately performed, and urine supernatant is frozen within few hours after collection. However, an adapted centrifuge (refrigerated and accommodating 50mL tubes) and an available technician might not be available at each clinical department or day-care hospital. Thus, valid information about necessity of an immediate centrifugation step is essential. Besides, if routinely collected, benchtop time of urine samples for chemokine quantification may vary. Here, we showed that uCXCL9 and uCXCL10 measurements where reproductible with storage up to 72H at RT, and without any prior centrifugation. This preanalytical study argues for a simple and feasible sample processing for clinical use: noninvasive collection of urine samples, no requirement for early centrifugation or PI addition, and stability at 4°C or RT up to 72H. Besides, if not available in all hospitals, urine samples could be shipped to a reference center for centralized quantification, or even home-collected. Indeed, telemedicine offers great potential to facilitate patient health care and to improve patient’s compliance to treatment. Home-based monitoring with remote self-collection could ensure a more comfortable and less stressful monitoring for patients.

To investigate the feasibility of clinical implementation of Ella® quantification for urine proteins, we compared ELISA and Ella® workflow, from sample thaw to render of the results. Upon assay preparation, Ella® appeared superior to conventional ELISA with no plate coating and no reagent preparation. Sample preparation only included thawing and an additional centrifugation step as to pellet all debris which might cause microfluidics and glass nanoreactors obstruction (**Figure 1, Assay preparation**). Ella® procedure further included a dilution step as for ELISA and a simple one-step sample deposition within the cartridge prior to running the assay (20 minutes). Once launched, time to result is approximately 70 minutes. Altogether, the estimated Ella® assay procedure is 1h30 as compared to 7 hours for a conventional ELISA (let alone antibody coating the day before for CXCL9/MIG DuoSet ELISA kit). Of importance, the Ella® assay only requires 35µL of urine supernatant to generate triplicate data, suggesting the possibility of quantifying more analytes from a single sample. Finally, most recent Ella® cartridges offer the possibility of a combined CXCL9/CXCL10 quantification, providing urine levels for both chemokines and for up to 32 samples (including low and high quality controls) within a fast turnaround time.

Both the ELISA and the Ella® method have inherent strengths and limitations. ELISA is a most validated and relative low-cost immunoassay, with universal availability. In comparison, Ella® technology appears less accessible, but with comparable price. Upon assay preparation, Ella® involved substantially less reagent preparation and clearly outperformed the ELISA in time-to-results. For both assays a number of constituents found in urine can interfere with their binding activity^26^; these compounds can vary from person-to-person based on differences in metabolism or even diet^27^. Because of this issue, one cannot designate either method as representing “truth.” We found the absolute values of uCXCL9 tended to be much higher (2.58 fold) when measured by the Ella® technique than when measured by ELISA. Although the absolute values of CXCL9 differed by laboratory method, results were very well correlated. Assessment of the ELISA kit using spiked concentration of recombinant CXCL9, suggested that it was not performing adequately in urine samples, for whom average percentage of expected values were poor (data not shown). Using the Ella® technology, recombinant CXCL9 levels were more consistently measured in urine matrix, with acceptable recovery within a urine-specific range of quantification. Overall, the relative low cost and ease of use of the fully-automated Ella® technology makes it amenable to the high laboratory throughput required in routine implementation.

After assessing that Ella® assay provided close results, in comparison to the reference ELISA technique, we assessed whether it provided useful results, *i.e.* similar diagnostic performance to assess the risk of AR. Although not identical, the high correlation between values from ELISA and Ella® ensured that individuals were ranked comparably by both methods, which is critical risk assessment models. Hence in cohort B, training the 8-parameter model on Ella®-data lead to similar accuracy, enabling the future building of a web application calculator for clinicians to use.

Our study has several limits. We individually addressed many parameters affecting urine sample storage. Given the inter-assay variation, the observed changes in uCXCL9 and uCXCL10 concentrations could be considered of minimal clinical relevance. However, only a storage on ice in a Styrofoam box for up to 72h could investigate real-life sample shipment conditions, where multiple factors may induce variability and affect reproducibility. Besides, though various storage conditions might be acceptable, within a single Transplant center or for an individual patient, keeping a standardized sample processing remains preferable. We acknowledge that our reproducibility measures for cohort A-D are not entirely comparable because the assays were performed several years apart. Thus, we cannot conclude whether these differences may arise from internal assay performance or from storage effects. Nevertheless, Ella® results are noninferior and could be even better in clinical use in which assays will be completed in a short period of time after collection.

In conclusion, the current study has identified and validated an improved method for the quantification of urine CXCL9 and CXCL10 for clinical use. Firstly, Ella® assay accurately measured both chemokines from urine samples. Secondly, a great simplification of preanalytical sample processing was reached with stability up to 72h at room temperature without any prior centrifugation or adjunction of preservative after urine collection. Thirdly, the fully-automated assay provides unprecedented rapid assessment, fitting the clinical expectations in render of the results. Fourthly, training our previously published model on Ella® data enabled to validate diagnostic accuracy and to develop an online available web application. Given these, urine CXCL9 and CXCL10 now display all characteristics for moving from research to clinical surveillance of AR in KTRs: time has come to put words into actions.

## Author Contributions

CT and DA conceived and designed the study. CL provided the urine samples, and XL and CN collected them. CT and LA collected the clinical data. VS, CN, CL, JAG and CT carried out the experiments. MR performed the histology reading of biopsies at Necker Hospital. CT, LS and LM performed the statistical analyses and interpreted the data. CL and FT participated in the interpretation of the results. CT and DA wrote the draft of the report. All the authors revised the report.

## Acknowledgments

We are indebted to all Necker Hospital teams and patients who participated in this research. We thank Xavier Lebreton, Joffrey Alves-Gasnier, Juliette Olivré and Lise Morin for their technical assistance. CT is supported by the *Société Francophone de Transplantation*. The Emmanuel Boussard Foundation supported DA, LA, LM, CT and VS, and DA received funds from the Day Solvay Foundation.

## Data availability statement

The data that support the findings of this study are available from the corresponding author upon reasonable request.

## ABBREVIATIONS

ABMR: antibody-mediated rejection
AR: acute rejection
AUC: area under the recipient operating curve
BKV: BK virus
CV: coefficient of variation
DSA: donor-specific antibody
ELISA: enzyme-linked immunosorbent assay
HLA: human leukocyte antigen
IQR: interquartile range
KD: kidney living-donors
KTR: kidney transplant recipient
LLOQ: lower limit of quantification
NPV: negative predictive value
PI: protease inhibitors
RT: room temperature
SD: standard deviation
uCXCL: urinary chemokine
ULOQ: upper limit of quantification

