## Supplemental Material for "Routine surveillance of kidney allograft rejection using a fully automated urinary CXCL9 and CXCL10 immunoassay"

**Supplemental Figure 1:**

ELISA quantification appeared to be a good option with robust results from our research laboratory experience, its availability in every hospital laboratory, and its affordable cost. However, the pilot chemokines study raised technical issues regarding the ELISA methods when used as a medium-throughput assay for urine chemokines quantification (unpublished data). CXCL10 was indeed quantified using a pre-coated ELISA plate within 6 hours, but CXCL9 quantification required an overnight coating of the plate prior to quantification, which proved difficult to transfer to routine. Next, we looked for alternate commercially available pre-coated ELISA kits quantifying CXCL9 and identified 3 candidates (**Supplemental Figure 1A**). We conducted a comparative test as to find which CXCL9 ELISA kit would be suitable for automatization and routine follow-up. Of importance, none of the ELISA kits had been validated on urine samples. All three ELISA kits produced very poor to uninterpretable results (**Supplemental Figure 1B**).

**Supplemental Table 1: Clinical and biological characteristics of kidney donors**

| **Clinical data** | | | **Laboratory results** | | | | | | | | | |
| --- | --- | --- | --- | --- | --- | --- | --- | --- | --- | --- | --- | --- |
|  |  |  | **uCXCL9 (pg/mL)** | | | **uCXCL10 (pg/mL)** | | | **Creatinine** | | **CBEU** | |
| **ID** | **Age** | **Sexe** | LOQ-censored | Uncensored quantification | uCXCL9/Cr (ng/mmol) | LOQ-censored | Uncensored quantification | uCXCL10/Cr (ng/mmol) | Serum Cr (µmol/L) | Urine Cr (mmol/L) | Leukocyturia (/mL) | Direct examination |
| **KD7*** | 31-35 | M | <LOQ | 0.0 | 0.0 | <LOQ | 0.8 | 0.1 | 82 | 15.8 | <1000 | No germ |
| **KD9** | 56-60 | F | <LOQ | 0.0 | 0.0 | <LOQ | 1.2 | 0.2 | 71 | 7.2 | 43400 | No germ |
| **KD5*** | 51-55 | F | <LOQ | 1.4 | 0.1 | <LOQ | 0.7 | 0.1 | 53 | 13.0 | 48600 | No germ |
| **KD4** | 56-60 | F | <LOQ | 0.4 | 0.1 | <LOQ | 0.5 | 0.2 | 68 | 3.1 | 1500 | No germ |
| **KD10** | 66-70 | M | <LOQ | 5.6 | 0.4 | 6.9 | 6.9 | 0.5 | 84 | 14.1 | 1700 | No germ |
| **KD3** | 36-40 | M | <LOQ | 15.1 | 0.5 | 9.4 | 9.4 | 0.3 | 72 | 30.9 | 15800 | No germ |
| **KD6** | 51-55 | F | <LOQ | 3.6 | 1.0 | 8.0 | 8.0 | 2.3 | 38 | 3.5 | 32400 | No germ |
| **KD1** | 31-35 | F | <LOQ | 27.5 | 1.3 | 13.0 | 13.0 | 0.6 | 58 | 20.8 | 6300 | No germ |
| **KD8*** | 66-70 | M | <LOQ | 6.6 | 2.4 | 2.1 | 2.1 | 0.8 | 75 | 2.7 | <1000 | No germ |
| **KD2** | 46-50 | F | <LOQ | 39.5 | 3.1 | 17.9 | 17.9 | 1.4 | 62 | 12.6 | 13400 | No germ |

Abbreviations: Cr, creatinine; CBEU, cytobacterial examination of the urine; F, female; ID, identity; KD, kidney donor; LOQ, limit of quantification; M, male. *Samples included in the recovery study. Uncensored quantification refers to non-validated values comprised between the limit of detection (LOD) and limit of quantification (LOQ).

**Supplemental Figure 1: Comparative assessment of 3 pre-coated CXCL9 ELISA kits**

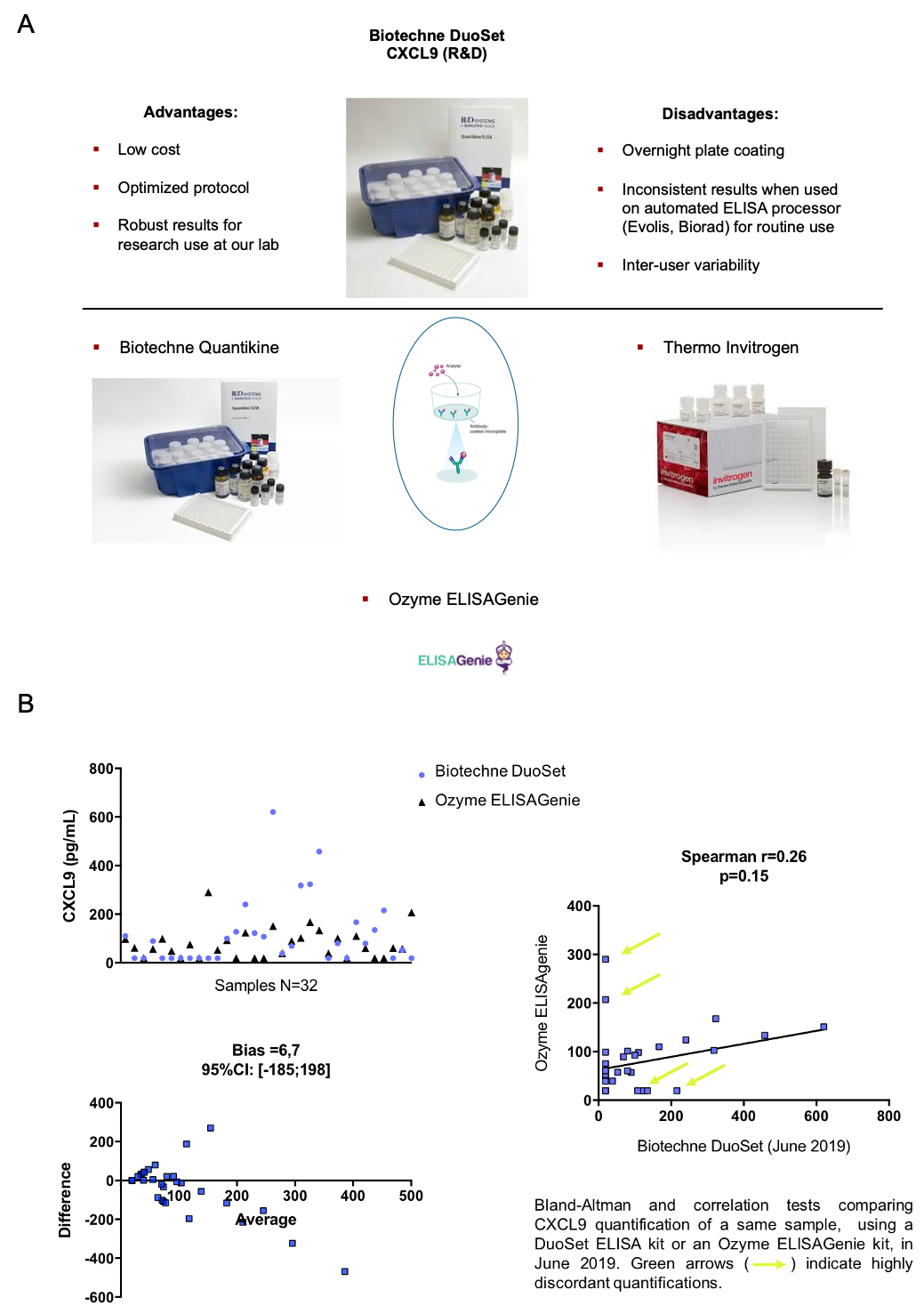
